# Thoracic expansion and spirometry 120 days after adding diaphragmatic manual therapy to physiotherapy in non-specific low back pain: Follow-up of a randomized controlled trial

**DOI:** 10.64898/2026.11.16.26368238

**Authors:** Francisco José Vera-Serrano, María Jesús Vinolo-Gil, Pedro Antonio Vera Serrano, Jesus Baltasar Gonzalez-Rubiño, Ismael García-Campanario, Juan Antonio Díaz-Mancha

**Affiliations:** Rehabilitation Clinical Management Unit, Interlevels-Intercenters Hospital Puerta del Mar, Hospital Puerto Real, Cádiz Bay-La Janda Health District, 11006 Cádiz, Spain; Research Unit, Biomedical Research and Innovation Institute of Cádiz (INiBICA), Puerta del Mar University Hospital, University of Cádiz, Ana de Viya 52, 11009 Cádiz, Spain; Department of Nursing and Physiotherapy, Faculty of Nursing and Physiotherapy, University of Cádiz, Ana de Viya 52, 11009 Cádiz, Spain; Rehabilitation Clinical Management Unit, Hospital Punta Europa, Campo de Gibraltar Oeste Health District, 11202 Algeciras, Cádiz, Spain; Department of Medicine, Faculty of Medicine, University of Cádiz, Cádiz, Spain; Physiotherapy Department, Faculty of Physiotherapy and Podiatry, University of Seville, C/Avicena s/n, 41009 Seville, Spain

## Abstract

**Objective:** An interim analysis of the DIAFRAGMA trial (46 participants) found that adding manual diaphragmatic stretching to physiotherapy improved thoracic expansion, forced vital capacity and forced expiratory volume in one second up to 30 days in non-specific low back pain. We assessed whether these differences persist at 120 days and, exploratorily, whether baseline values moderate the response.

**Methods:** In this parallel-group, assessor-blinded randomized controlled trial, 55 adults were randomized and 50 were analysed (22 experimental, 28 control). All participants received ten sessions of conventional physiotherapy; the experimental group also received manual diaphragmatic stretching in six of them. Thoracic expansion (cirtometry at three levels) and portable spirometry were assessed at baseline, after the first and tenth sessions, and 7, 30 and 120 days after treatment. Between-group differences were estimated with linear mixed-effects models and false discovery rate correction.

**Results:** At 120 days, inspiratory thoracic expansion was greater in the experimental group at the axillary (3.38 cm; 95% CI 2.94 to 3.81), xiphoid (3.52 cm; 2.78 to 4.26) and basal levels (4.12 cm; 3.36 to 4.88). Forced vital capacity (1.57 L; 0.96 to 2.18), forced expiratory volume in one second (1.49 L; 0.97 to 2.01) and peak expiratory flow (2.70 L/s; 1.21 to 4.19) were also higher (all p < 0.001). These differences arose during treatment and remained stable thereafter. Expiratory-phase expansion and mid-to-late expiratory flows did not improve. No baseline value moderated the response.

**Conclusions:** Adding manual diaphragmatic stretching to physiotherapy was associated with greater inspiratory chest expansion and higher spirometric values that were maintained at 120 days. Because of the size of the spirometric differences, the portable spirometry protocol and the lack of participant blinding and a sham comparator, these findings require confirmation in sham-controlled trials using laboratory spirometry.

**Trial registration:** ClinicalTrials.gov NCT06069388.

## 1. Introduction

Low back pain is the leading cause of years lived with disability worldwide. Global Burden of Disease estimates indicate that approximately 619 million people were affected by low back pain in 2020, projected to reach 843 million by 2050 [1], and low back pain a contributor to years lived with disability across age groups [2]. A substantial proportion of these cases are classified as non-specific, with no identifiable structural cause, which has progressively shifted the therapeutic focus from pain management alone towards restoring function [3–5].

Among the structures implicated in this functional approach, the diaphragm has attracted growing attention. Beyond its primary role in ventilation, the diaphragm contributes to intra-abdominal pressure regulation and lumbar spine stabilisation through its crural insert on the upper lumbar vertebrae, and it is anatomically connected with the thoracolumbar fascia, psoas major and quadratus lumborum [6,7]. Diaphragmatic function has been linked to postural control and balance [8], and physiological studies show that the diaphragm is recruited during postural tasks independently of its respiratory rhythm, with its postural coordination differing between people with and without chronic low back pain [9,10]. This dual respiratory and postural role makes the diaphragm a plausible target for physiotherapy in NSLBP and a rationale for measuring chest wall mobility and lung function. Chronic low back pain extends beyond isolated musculoskeletal pathology, directly involving ventilatory biomechanics and motor control. The diaphragm and deep abdominal muscles perform an essential dual function: respiration and postural stabilization of the trunk. In patients with low back pain, it has been extensively documented that this shared physiological function is compromised; this manifests as altered diaphragmatic positioning and kinematics—compensating for a lack of lumbopelvic stability [10] —alongside increased fatigability of the inspiratory muscles and a high prevalence of dysfunctional breathing patterns [11]. In these individuals, reductions in parameters such as forced vital capacity (FVC) or maximal voluntary ventilation (MVV) do not reflect structural damage to the lung parenchyma or intrinsic airway obstruction; rather, they serve as direct markers of profound neuromuscular and biomechanical dysfunction [12]. Therefore, altered spirometric parameters in low back pain reveal the extent to which impaired core stabilization has affected the respiratory bellows [13]. Identifying these functional changes demonstrates that the management of low back pain should not be limited to the spine; instead, it requires re-education of breathing patterns and inspiratory muscle training to restore the synergy between respiration and postural stability.

Several small trials evaluating manual techniques applied directly to the diaphragm have reported short-term improvements in thoracic mobility, spirometric parameters or pain [14–17]. The DIAFRAGMA trial (ClinicalTrials.gov NCT06069388) tested whether adding a manual diaphragm-stretching technique, delivered in six of ten sessions, to conventional physiotherapy improves thoracic expansion and pulmonary function in NSLBP. A first report of 46 participants (20 experimental, 26 control) found greater basal thoracic expansion, FVC and FEV1 after treatment, maintained at 30 days [18].

That interim report explicitly identified the absence of long-term follow-up as a limitation preventing any assessment of the durability of these effects beyond one month [18]. This is not a limitation specific to this trial: a 2026 umbrella review of manual therapy for chronic NSLBP concluded that methodological quality and short follow-up across the included evidence base constrain any conclusion about long-term efficacy, and that treatment effects for this condition more commonly diminish rather than persist over time [19].

The first report also covered only basal inspiratory expansion, FVC and FEV1, leaving unreported the axillary- and xiphoid-level expansion coefficients, the expiratory phase of chest wall mobility, and peak expiratory flow (PEF) and the mid-to-late expiratory flow indices (FEF25%, FEF75% and FEF25–75%) that were collected under the same protocol but not reported in that initial publication. The clinical value of FEF25–75% and FEF75% beyond FEV1 is often debated because of their large between-subject variability [20]. Furthermore, in a recent systematic review protocol published by our research group [21], we specifically identified both the lack of long-term follow-up and the absence of standardized reporting for respiratory outcomes as critical evidence gaps in this field.

We therefore analyzed the complete DIAFRAGMA cohort (n=50) to determine, first, whether the differences reported at 30 days [18] persist at 120 days; second, whether comparable differences exist at 120 days for the previously unreported expansion levels, expiratory phase and flow indices; and third, as an exploratory aim, whether the baseline value of each outcome moderates the 120-day response. We hypothesized *a priori* that the differences observed at 30 days would persist at 120 days, and that comparable improvements would be found for the newly analyzed respiratory variables. Regarding the exploratory aim, we postulated that the baseline status could modify the treatment response, as patients with more restricted initial mobility or worse pulmonary function might have a greater margin for improvement.

## 2. Materials and Methods

### 2.1. Study design, registration and relationship to the interim report

This report presents the 120-day follow-up of the DIAFRAGMA trial, a parallel-group randomized, assessor-blinded controlled trial registered at ClinicalTrials.gov (NCT06069388; first submitted September 18, 2023 and approved by the Research Ethics Committee of the University of Seville (ref. 1032-N-23). The registered primary outcome was basal inspiratory expansion assessed at 30 days [18]. The four-month (120-day) assessment time point and all respiratory outcomes reported in this article—including expansion levels, the expiratory phase, and flow indices—were explicitly pre-specified in the registered protocol. The trial registry has been updated to reflect its ‘completed’ status and the actual study dates. The report follows the CONSORT 2025 statement (S1 CONSORT Checklist).

The first report [18] analyzed 46 participants (20 experimental, 26 control) in a data cut that was not based on a pre-specified interim analysis or stopping rule. The data cut-off date for that preliminary report was in October 2024. Although trial recruitment ended in September 2024, the final four participants had not yet completed their 30-day assessment by the time of that initial data extraction, which explains their exclusion from the earlier publication. The present analysis includes the complete cohort of 50 participants (22 experimental, 28 control). The last visit of the last participant occurred on January 2025, and the final database lock was completed on February 2025. Where the two reports overlap (basal inspiratory expansion, FVC and FEV1 up to 30 days), the estimates presented here supersede those of [18]; they differ because the sample is larger and because a different statistical model was used (linear mixed-effects models in the present study instead of the repeated-measures ANOVA used in [18].

The study was approved by the Research Ethics Committee of the University of Seville, Spain (ref. 1032-N-23) and was conducted in accordance with the Declaration of Helsinki. All participants provided written informed consent before enrolment. The trial protocol approved by the ethics committee is provided as S1 Protocol, and the completed CONSORT 2025 checklist as S1 CONSORT Checklist. A related cross-sectional analysis of the baseline data of this cohort has been published separately [22]. The protocol approved by the ethics committee (S1 Protocol) differs from the conduct and analysis of the trial in the following respects. (1) The protocol did not designate a primary outcome. (2) The protocol planned repeated-measures analysis of variance; linear mixed-effects models with false discovery rate correction were used instead to model all six assessments with a participant-level random effect, and the moderator and sensitivity analyses were not prespecified. (4) Handling of missing data and adverse-event monitoring were not specified in the protocol.

### 2.2. Participants, randomization and blinding

Recruitment took place at a private physiotherapy clinic in Chiclana de la Frontera (Cádiz, Spain) between December 2023 and September 2024 [18]. The source population consisted of patients referred through a private health insurance network; we ensured that no participant had an open compensation claim or active litigation related to their back pain. Eligible participants were adults aged 18 years or older with a specialist diagnosis of subacute or chronic NSLBP (ICD-10 M54.5)—defined as symptoms lasting between 4 and 12 weeks for subacute, or more than 12 weeks for chronic—and diaphragm dysfunction (≥2 of: apical breathing pattern, accessory muscle overactivity at rest, breath-holding time <15 s, and restricted costal mobility) [23–25]. Exclusion criteria were specific structural spinal pathology, recent spinal or thoracoabdominal surgery, systemic or neurological conditions affecting the lumbar spine or diaphragm, pre-existing primary respiratory diseases (such as asthma or COPD), and previous diaphragmatic manual treatment, among others listed in [18].

Of 64 people assessed for eligibility, 9 were excluded because they could not attend ten consecutive sessions, and 55 were randomized (27 to the experimental group, 28 to the control group). Five participants allocated to the experimental group withdrew after randomization and before the baseline assessment (work commitments, n=1; travel, n=1; declined participation, n=3) and contributed no data. One control participant discontinued after session 1, and the last observed values were carried forward (LOCF) for all later assessments. Fifty participants (22 experimental, 28 control) were analyzed, 49 of them with observed data at all six assessments (Fig 1). There was no patient or public involvement in the design, conduct or reporting of the trial.

**Fig 1.**
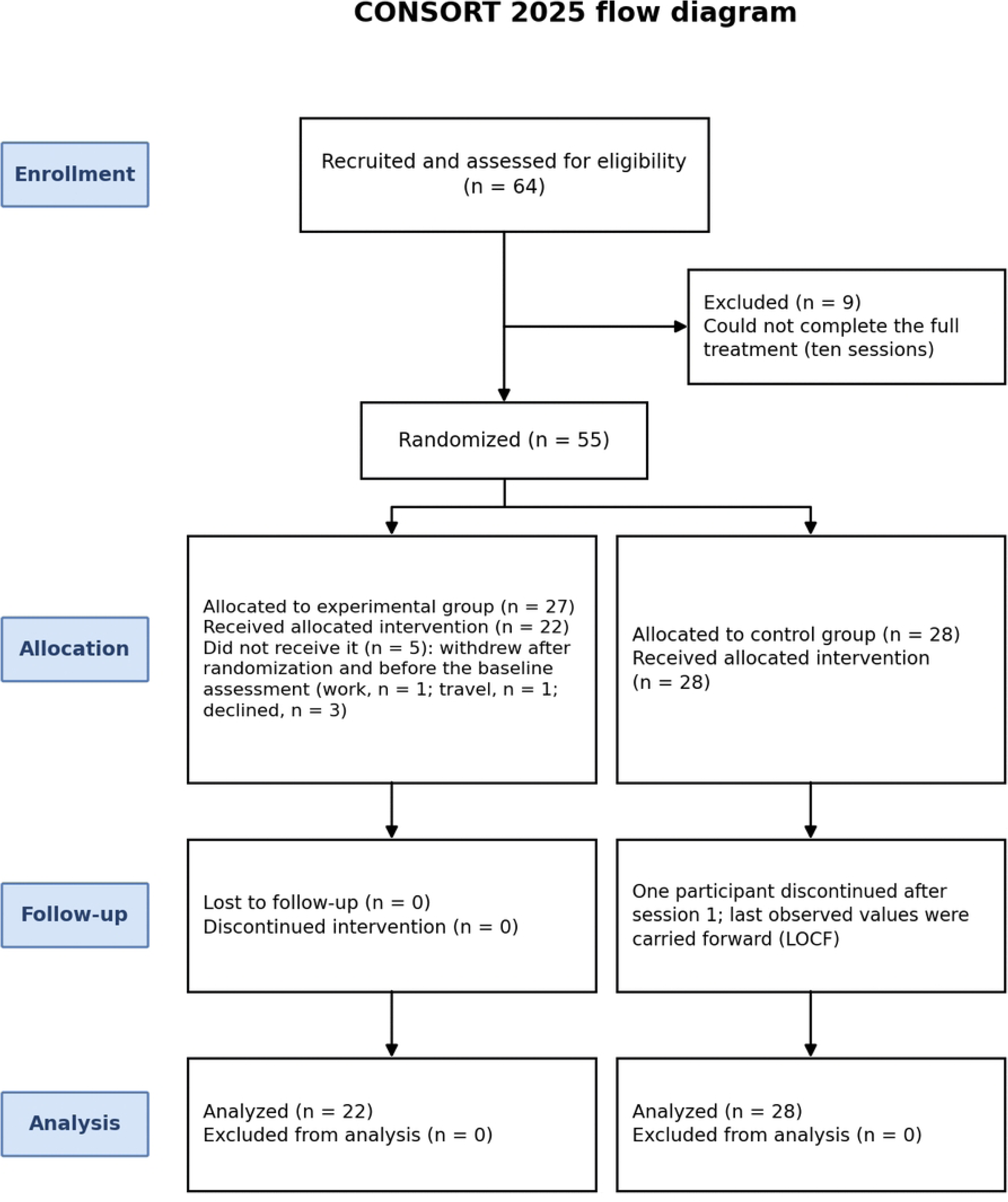
CONSORT 2025 participant flow diagram. The control participant who discontinued after session 1 was retained in the analysis using last observation carried forward for subsequent assessments; therefore, the number analysed equals the number allocated in that group.

Regarding trial methodology details previously reported [18], the sample size was calculated to ensure 90% power to detect a clinically meaningful difference in the primary outcome. An independent researcher generated the simple randomization sequence using computer-generated random numbers. Allocation was concealed using sequentially numbered, opaque, sealed envelopes, and only the unblinded treating physiotherapist had access to the sequence. Crucially, all baseline assessments were conducted strictly after randomization. Both the outcome assessor and the data analyst were blinded to group allocation; this blinding was maintained by explicitly instructing participants not to disclose their treatment and by masking group identifiers in the final database.

### 2.3. Intervention

Both groups received standard physiotherapy consisting of tetrapolar transcutaneous electrical nerve stimulation, infrared lamp therapy, therapeutic ultrasound over the quadratus lumborum, and ischemic compression with analytical stretching of the quadratus lumborum, multifidus and iliocostalis muscles, delivered across ten daily sessions, as previously described [18]. At the end of six sessions (1, 3, 5, 6, 8 and 10), the experimental group also received manual diaphragmatic stretching technique. Supported by established manual therapy frameworks [26–28]: the therapist placed the ulnar borders of both hands on the lower costal margin (approximately ribs 8 to 10) with the patient supine and a cushion under the knees. The therapist applied bilateral cephalad traction at the beginning of deep inspiration, maintained it at peak inspiration and throughout expiration, and kept verbal contact with the patient for ten breathing cycles (about 3 to 5 minutes). A brief guide to diaphragmatic breathing was provided strictly on an-as-needed basis to those specific participants who had difficulty initiating deep breaths on demand; it was not routinely administered to the entire cohort. All treatments were delivered by one physiotherapist, with a minimum of 15 years of clinical experience. To ensure intervention fidelity and consistency, the therapist completed a 50-hour standardization workshop prior to the trial, and the exact same personnel treated participants in both the experimental and control groups. The remaining four sessions of the experimental group and all ten control sessions comprised standard physiotherapy only. While no home exercise program or other care was prescribed during the trial or the 120-day follow-up, participants were systematically asked about the use of any non-prescribed co-interventions (e.g., analgesic use or independent exercise routines) at each assessment, and these were prospectively recorded. A treatment manual and completed TIDieR checklist [29] are available as S1 Appendix.

### 2.4. Outcome measures

All outcomes were assessed at baseline, immediately after the first and tenth sessions, and 7, 30 and 120 days after the last session. All measurements across all time points were performed by a single blinded assessor who had completed a specific 20-hour standardization training program prior to the trial.

Thoracic wall expansion was measured by cirtometry with a non-elastic tape at three standardised anatomical levels — axillary, xiphoid, and basal — with the patient standing, performing maximal inspiration and maximal expiration. The amplitude coefficient at each level was calculated as previously described [18,30], and, unlike the primary report (which considered only inspiratory expansion at the basal level), both the inspiratory and expiratory phases at all three levels are analyzed here. Spirometric parameters (FVC, FEV1, PEF, FEF25%, FEF75% and FEF25–75%) were obtained with a portable spirometer (CONTEC SP10W, Contec Medical Systems, Qinhuangdao, China) using the protocol described in the interim report [18]. Device calibration was checked before each testing session following American Thoracic Society recommendations. With the participant seated, each manoeuvre consisted of a maximal inspiration followed by a forced expiration lasting at least 1.5 s; at least two manoeuvres were performed, separated by one-minute rest intervals, and the mean of the two manoeuvres was recorded [31]. FVC values in this study therefore represent the volume exhaled during a short forced manoeuvre on a portable device, which depends on participant effort and technique. Only FVC and FEV1 were reported in the interim publication [18]. Adverse events were monitored by the treating physiotherapist and the outcome assessor at every session and assessment. In the interim report, mild transient muscular discomfort and fatigue were reported by one participant in the experimental group and two in the control group, all resolving without intervention [18]. No further adverse events were recorded through the 120-day follow-up.

### 2.5. Statistical analysis

Continuous variables are described as mean (standard deviation); values by group and time point are given in S1 Table. Between-group differences over time for each outcome were modeled using linear mixed-effects models with group, time and the group × time interaction as fixed effects and a random intercept per participant, fit by restricted maximum likelihood. All 50 analysed participants were included in every model (modified intention-to-treat; see 2.2 for the single LOCF imputation). Estimated between-group differences with 95% confidence intervals are reported at each post-baseline time point; the estimates for basal inspiratory expansion, FVC and FEV1 up to 30 days are re-estimated here, in the complete cohort, for context alongside the new 120-day time point (see 2.1). Given the acknowledged measurement variability and debated clinical interpretability of FEF25%, FEF75% and FEF25–75% [13], these three indices were treated as exploratory. Within each outcome family (thoracic expansion, six variables; spirometry, six variables), p-values across all post-baseline time points were corrected for multiple comparisons using the Benjamini-Hochberg false discovery rate procedure [32]; both uncorrected and FDR-corrected p-values are reported, and statistical significance was defined as FDR-corrected *p*<0.05. To assess the robustness of the 120-day estimates, we performed two sensitivity analyses. First, the 120-day value of each outcome was compared between groups using analysis of covariance with the baseline value as covariate. Exact uncorrected and FDR-corrected p-values for all comparisons are given in S2 Table, and full regression results of the moderator analysis in S3 Table. Analyses were performed with R version 4.6.2 with package lme4. A two-sided alpha of 0.05 was used.

### 2.6. Moderator analysis

As an exploratory secondary aim, we tested whether a participant’s baseline value on each outcome moderated the magnitude of their 120-day change. For each of the 12 outcomes, change from baseline to 120 days was regressed on group, mean-centered baseline value, and their interaction; the group × baseline interaction term tests whether baseline severity differentially predicts the treatment effect (moderation), as distinct from simple within-group regression to the mean [33]. This analysis was limited to the outcome’s own baseline value, since demographic and clinical covariates beyond age, sex, height, weight and BMI (Table 1) were not available for this analysis; a fuller moderator analysis incorporating symptom chronicity, if available, would be a natural extension. Interaction p-values across the 12 outcomes were FDR-corrected.

**Table 1.** Demographic and anthropometric characteristics of the study sample.

| **Variable** | **Experimental (n=22)** | **Control (n=28)** | **p** |
|---|---|---|---|
| Age, years, mean (SD) [range] | 43.82 (15.10) [23-72] | 47.64 (16.87) [18-74] | .36 |
| Sex: female, n (%) | 9 (40.9%) | 13 (46.4%) | .70 |
| Height, cm, mean (SD) | 172.46 (8.08) | 168.79 (9.56) | .14 |
| Weight, kg, mean (SD) | 81.37 (14.68) | 76.58 (19.84) | .31 |
| BMI, kg/m², mean (SD) | 27.32 (4.26) | 26.70 (5.82) | .64 |

*Baseline demographic and anthropometric characteristics. Between-group comparisons by independent-samples t-test or Mann–Whitney U test as appropriate. No significant differences were found for any variable. EG: experimental group; CG: control group; SD: standard deviation; BMI: body mass index.*

## 3. Results

### 3.1. Participants

Fifty participants (22 experimental, 28 control) were analyzed; 49 had observed data at all six assessments and one control participant contributed data carried forward from session 1 (Fig 1). This is the same baseline cohort analyzed in a related cross-sectional report from this trial [34], confirmed by the matching sample size (N=50) and sex distribution (22 women, 28 men). The experimental group had higher baseline values for the expiratory flow indices (PEF, FEF25%, FEF75% and FEF25–75% (Table 2) No adverse events related to either intervention were reported throughout the trial.

**Table 2.**
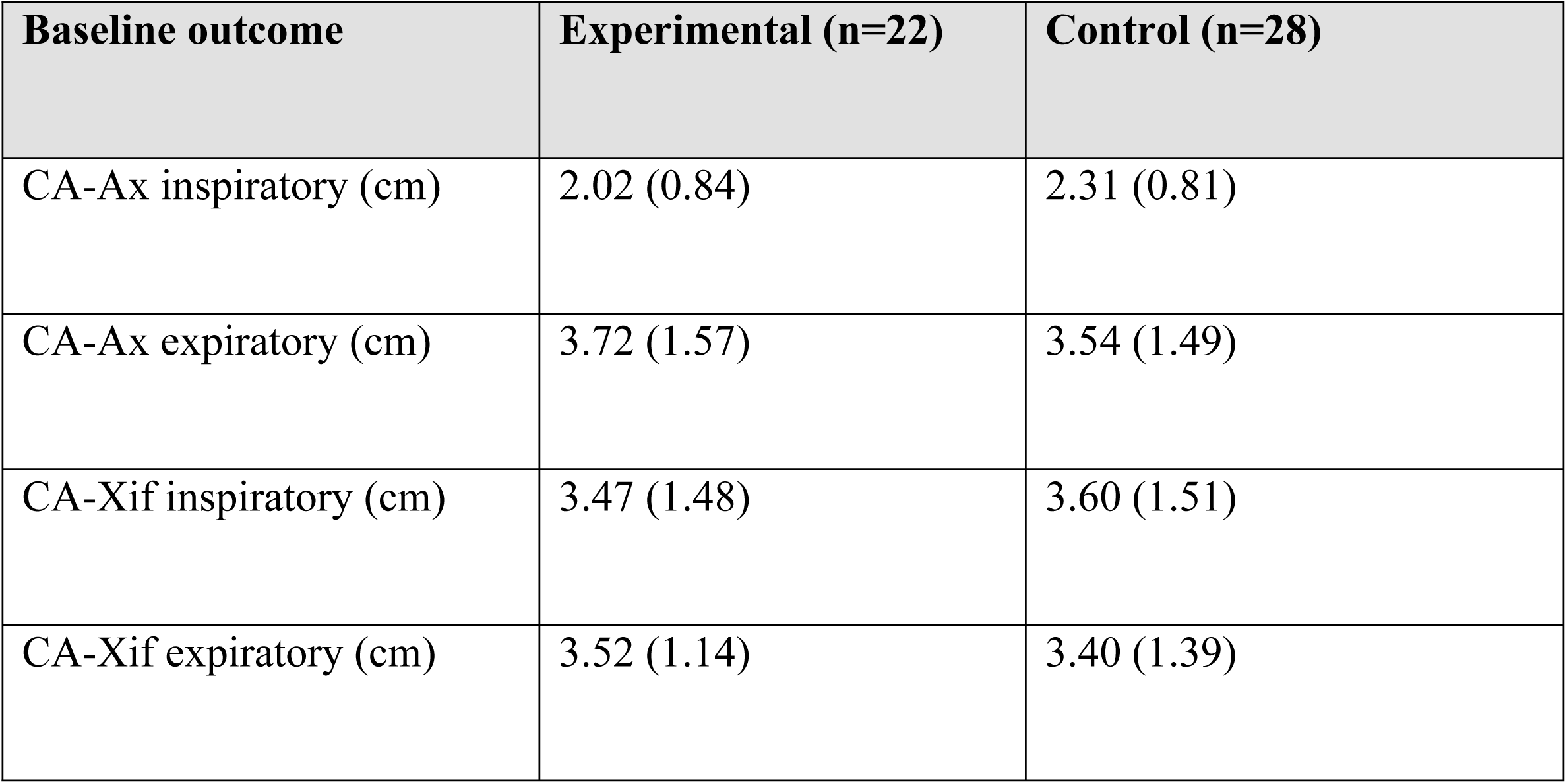
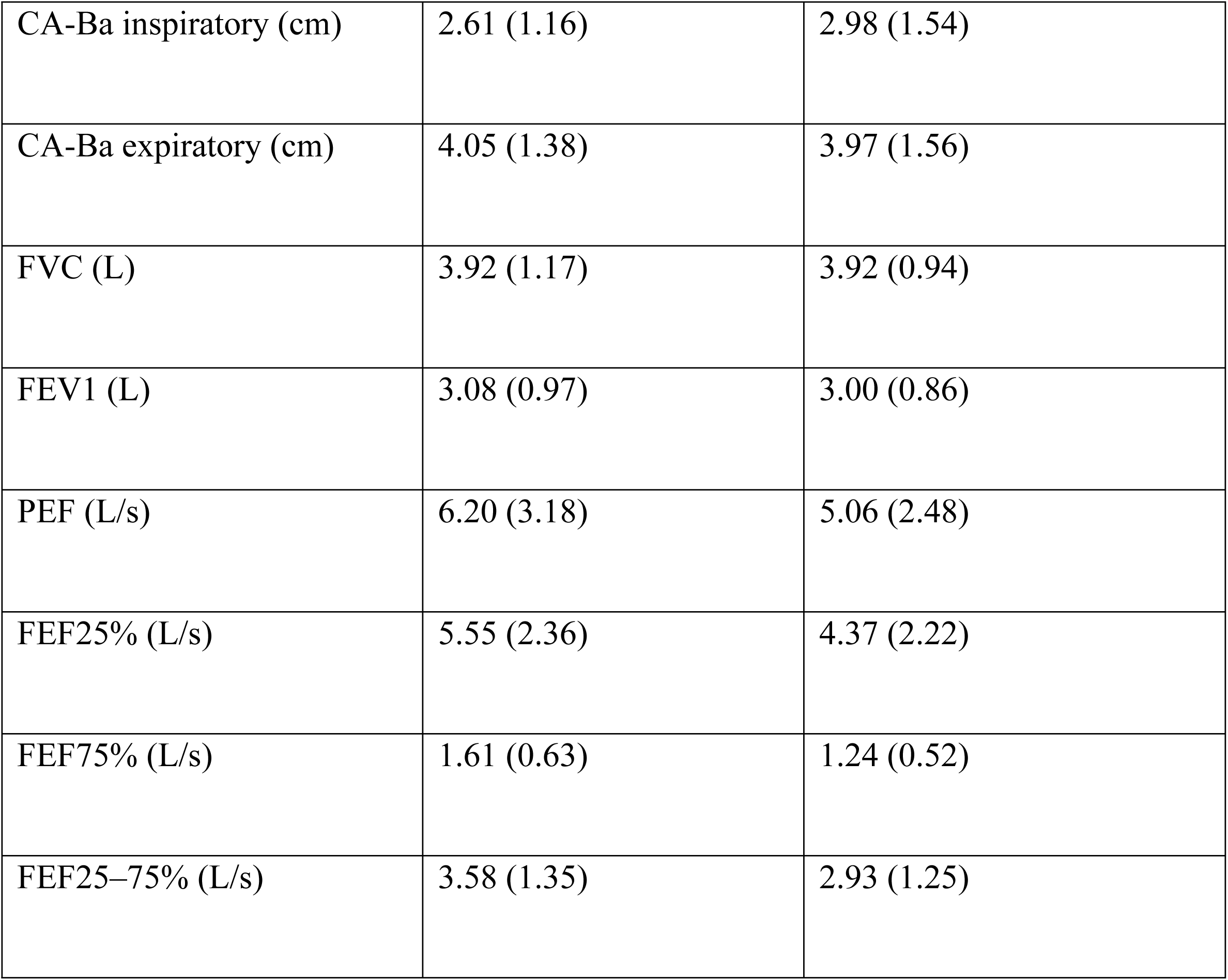
Baseline characteristics of the participants. Values are mean (SD) unless otherwise stated. CA-Ax, CA-Xif and CA-Ba: amplitude coefficient at the axillary, xiphoid and basal levels.

### 3.2. Thoracic wall expansion

Inspiratory expansion increased progressively in the experimental group relative to control at all three anatomical levels, reaching its largest between-group difference at 120 days for every level (Table 3). Expiratory-phase expansion showed no between-group benefit at any level; at the xiphoid and basal levels it numerically favored the control group from day 7 onward, reaching statistical significance at several time points (marked * in Table 3, surviving FDR correction). From baseline to 120 days, inspiratory expansion increased in the experimental group (axillary +2.77 cm, xiphoid +2.64 cm, basal +3.60 cm) and decreased in the control group (−0.88, −1.01 and −0.88 cm; S1 Table).

**Table 3.** Between-Group Comparisons by Outcome and Follow-up Point thoracic wall expansion.

| **Variable** | **T2 Post-session 1** | **T3 Post-session 10** | **T4 Day 7** | **T5 Day 30** | **T6 Day 120** |
|---|---|---|---|---|---|
| CA-Ax inspiratory | **1.27 [0.83, 1.70]\*** | **2.93 [2.49, 3.37]\*** | **3.00 [2.56, 3.43]\*** | **2.88 [2.44, 3.31]\*** | **3.38 [2.94, 3.81]\*** |
| CA-Ax expiratory | 0.01 [-0.61, 0.64] | -0.48 [-1.10, 0.14] | -0.45 [-1.07, 0.18] | -0.20 [-0.82, 0.42] | -0.39 [-1.01, 0.23] |
| CA-Xif inspiratory | **1.50 [0.75, 2.24]\*** | **3.13 [2.39, 3.87]\*** | **3.11 [2.37, 3.85]\*** | **3.17 [2.42, 3.91]\*** | **3.52 [2.78, 4.26]\*** |
| CA-Xif expiratory | -0.33 [-0.95, 0.28] | -0.39 [-1.01, 0.22] | **-0.72 [-1.33, -0.10]\*** | **-0.74 [-1.36, -0.12]\*** | **-0.87 [-1.49, -0.25]\*** |
| CA-Ba inspiratory † | **2.03 [1.26, 2.79]\*** | **3.70 [2.94, 4.46]\*** | **3.80 [3.03, 4.56]\*** | **3.90 [3.14, 4.66]\*** | **4.12 [3.36, 4.88]\*** |
| CA-Ba expiratory | -0.03 [-0.71, 0.65] | **-0.75 [-1.43, -0.07]\*** | -0.67 [-1.34, 0.01] | **-0.81 [-1.49, -0.13]\*** | **-1.20 [-1.88, -0.53]\*** |

*Between-group mean differences in cm (experimental minus control) from linear mixed-effects models, with 95% CI. \* FDR-corrected p<.05. † Outcome partially reported to 30 days in the interim trial report [11]; re-estimated here in the complete cohort for context. Exact uncorrected and FDR-corrected p-values for all comparisons are given in S2 Table.*

### 3.3. Spirometric parameters

Between-group differences in FVC, FEV1 and PEF emerged during the treatment period and remained stable through 120 days; all survived FDR correction at every post-baseline time point except FVC immediately after the first session (Table 4). In the experimental group, mean FVC increased by 0.30 L after the first session and by a further 0.89 L between the first and tenth sessions, and changed by only 0.04 L between the tenth session and day 120. In the control group, FVC decreased progressively (−0.12, −0.05 and −0.14 L over the same intervals). From baseline to 120 days, FVC increased by 1.23 L in the experimental group and decreased by 0.30 L in the control group; FEV1 increased by 1.21 L and decreased by 0.17 L; and PEF increased by 1.44 L/s and decreased by 0.11 L/s (S1 Table). FEF25% and FEF75% showed nominal differences after the first session that did not survive FDR correction; FEF25–75% showed no between-group difference at any time point.

**Table 4.** Between-Group Comparisons by Outcome and Follow-up Point spirometric parameters.

| **Variable** | **T2 Post-session 1** | **T3 Post-session 10** | **T4 Day 7** | **T5 Day 30** | **T6 Day 120** |
|---|---|---|---|---|---|
| FVC (L) † | 0.46 [-0.15, 1.07] | **1.40 [0.79, 2.01]\*** | **1.41 [0.80, 2.02]\*** | **1.46 [0.85, 2.07]\*** | **1.57 [0.96, 2.18]\*** |
| FEV1 (L) † | **0.72 [0.21, 1.24]\*** | **1.41 [0.89, 1.93]\*** | **1.45 [0.93, 1.97]\*** | **1.51 [0.99, 2.03]\*** | **1.49 [0.97, 2.01]\*** |
| PEF (L/s) | **2.03 [0.54, 3.52]\*** | **2.27 [0.78, 3.76]\*** | **2.36 [0.87, 3.85]\*** | **2.52 [1.03, 4.01]\*** | **2.70 [1.21, 4.19]\*** |
| FEF25% (L/s) | 1.27 [0.13, 2.40] | 0.85 [-0.29, 1.99] | 0.77 [-0.37, 1.91] | 1.14 [0.00, 2.28] | 1.16 [0.02, 2.30] |
| FEF75% (L/s) | 0.01 [-0.39, 0.42] | 0.11 [-0.29, 0.51] | 0.09 [-0.31, 0.49] | 0.18 [-0.22, 0.59] | 0.22 [-0.18, 0.62] |
| FEF25–75% (L/s) | 0.66 [-0.25, 1.56] | -0.02 [-0.92, 0.88] | 0.71 [-0.19, 1.61] | 0.28 [-0.63, 1.18] | 0.61 [-0.30, 1.51] |

*Between-group mean differences from linear mixed-effects models, with 95% CI. \* FDR-corrected p<.05. † Outcome partially reported to 30 days in the interim trial report [11]; re-estimated here in the complete cohort for context. Exact uncorrected and FDR-corrected p-values for all comparisons are given in S2 Table.*

The direction of change was highly consistent at the individual level: 21 of 22 participants in the experimental group and none of 28 in the control group showed an FVC gain of at least 0.20 L and 12% at 120 days, whereas 16 of 28 control participants showed a decrease of 0.15 L or more. Baseline-adjusted analysis of covariance gave 120-day differences of 1.53 L (95% CI 1.33 to 1.73) for FVC and 1.39 L (95% CI 1.16 to 1.63) for FEV1, consistent with the mixed-model estimates (S4 Table). Because baseline PEF was higher in the experimental group, the baseline-adjusted 120-day difference for PEF was smaller than the unadjusted estimate (1.79 L/s; 95% CI 0.98 to 2.61).

Randomization balance was adequate for 11 of 12 outcomes at baseline (all FDR-corrected *p*>.05); FEF25% showed a nominally significant baseline imbalance favoring the experimental group (raw p=0.043), so between-group estimates for FEF25% at follow-up should be interpreted with added caution.

### 3.4. Correlations between thoracic expansion and spirometry at 120 days

At 120 days, inspiratory expansion at all three levels correlated strongly with both FVC and FEV1 (basal: rho=0.71 with FVC, rho=0.74 with FEV1; axillary: rho=.67 with FVC, rho=0.71 with FEV1; xiphoid: rho=0.72 with FVC, rho=0.75 with FEV1; all p<0.001, n=50), a pattern of comparable or slightly greater strength than the correlations reported at 30 days in the interim report (rho=0.73 with FVC, rho=0.75 with FEV1, basal level only) [18], indicating that the association between chest wall mobility and pulmonary function is maintained rather than weakening over the long term.

### 3.5. Moderators of the 120-day response

No baseline value moderated the magnitude of 120-day change for any of the 12 outcomes after correction for multiple comparisons (Table 5). A nominally significant interaction was observed for FVC (uncorrected *p*=0.012), suggesting participants with higher baseline FVC may have shown a larger treatment-related gain, but this did not survive FDR correction (*p*=0.142) and should be treated as hypothesis-generating only.

**Table 5.**
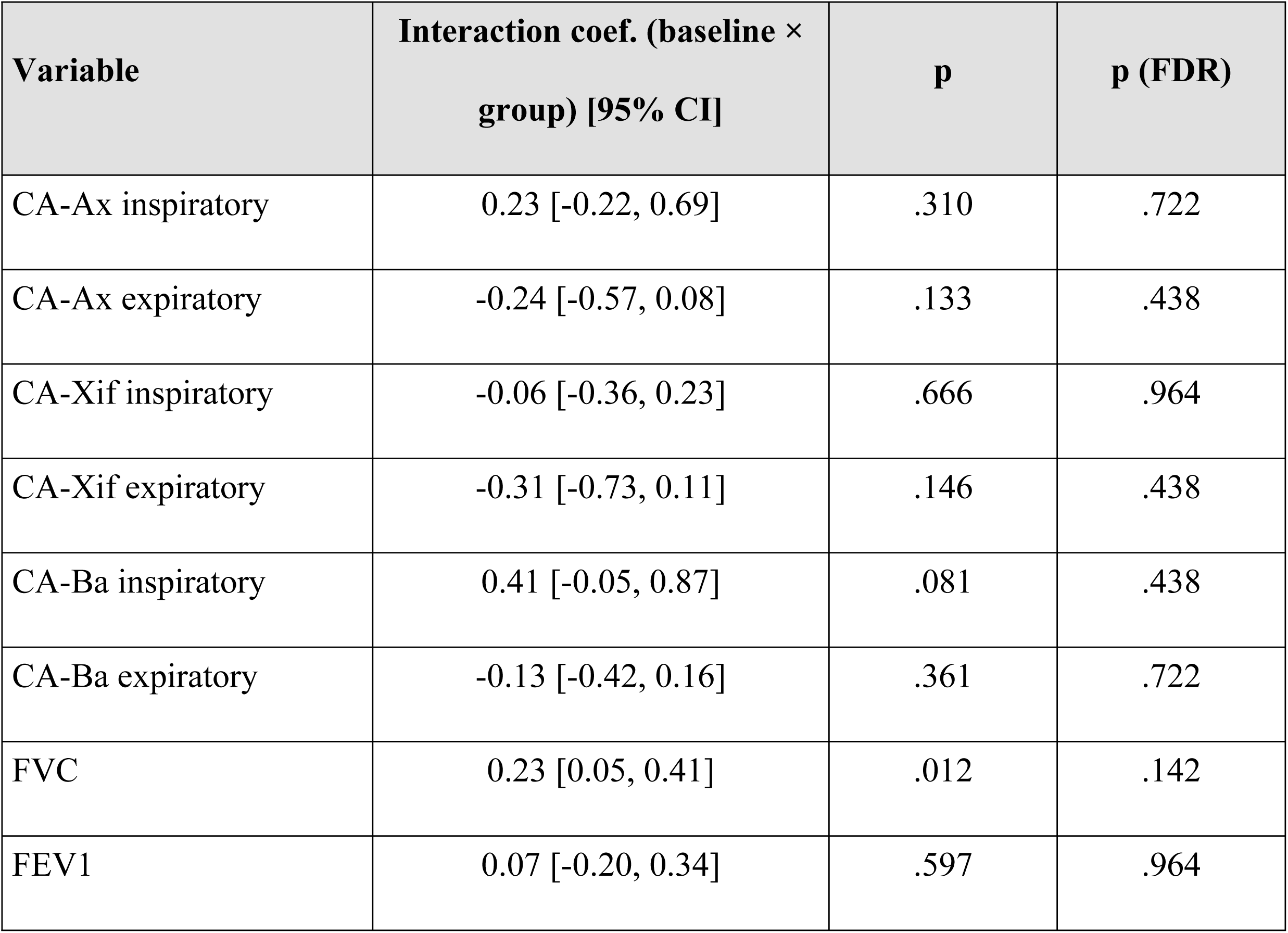
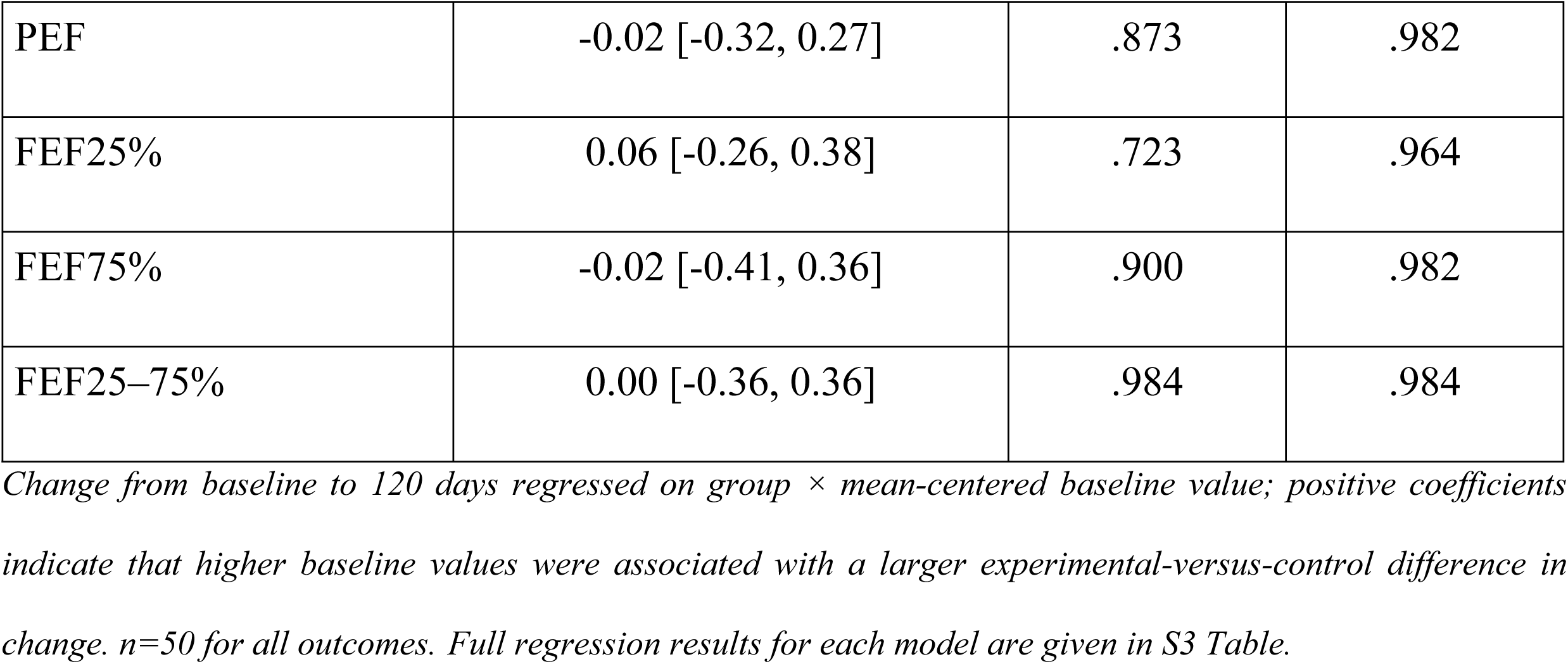
Exploratory moderator analysis of the 120-day response: group × baseline-value interaction on the change from baseline to 120 days.

## 4. Discussion

In the complete DIAFRAGMA cohort, the between-group differences in inspiratory thoracic expansion, FVC and FEV1 previously reported to 30 days in an interim analysis [18] were maintained at 120 days. The differences arose during the ten-session treatment period and remained stable thereafter, while values in the control group declined slightly over the same period. The same pattern extended to the axillary and xiphoid expansion levels and to peak expiratory flow, none of which had previously been reported for this trial. Maintenance of the difference contrasts with the broader manual therapy literature for chronic low back pain, where treatment effects more typically diminish over time [19]; whether this reflects a feature of diaphragm-targeted techniques or of the respiratory and mechanical outcomes examined here, rather than pain-based outcomes, cannot be determined from a single trial. A lasting change in the mechanical or fascial properties of the diaphragm and chest wall has been proposed for these techniques [6,7].

The magnitude of the spirometric differences requires cautious interpretation. A mean FVC increase of 1.23 L (about 33% of baseline) in adults without respiratory disease is far larger than the 100–200 mL changes usually considered clinically important [18] and the control group also showed a decline in FVC of about 0.3 L over four months. Some features of the trial may have contributed. First, participants were not blinded, and those in the experimental group received additional hands-on contact and, when needed, guidance on diaphragmatic breathing, which may have improved their performance of the manoeuvre (a learning effect), while control participants may have become less motivated over repeated testing. Second, a single assessor performed all measurements. For these reasons, the spirometric findings should be read as changes in the volume exhaled during a short forced manoeuvre on a portable device, and they need confirmation with laboratory spirometry in a sham-controlled trial.

The picture was materially different for expiratory-phase mobility, which showed no benefit and, at the xiphoid and basal levels, numerically favored the control group from day 7 onward. This asymmetry between inspiratory and expiratory phases is consistent with the diaphragm’s primary biomechanical role in inspiration, and mirrors the interim report’s own observation that expiratory expansion did not change at the basal level [18]; the present results extend that observation to all three levels and to a much longer follow-up, and additionally suggest the difference may not be a null finding so much as a genuine divergence between phases that merits its own mechanistic explanation, which the present data cannot provide. Similarly, the null or fragile findings for FEF25%, FEF75% and FEF25–75% should not be read as evidence against the intervention: these indices carry substantial between-subject measurement variability, and their incremental clinical value beyond FEV1 has been directly questioned in the pulmonary function literature [20]. FEF25% additionally showed a nominal baseline imbalance between groups, further limiting confidence in any isolated significant finding for that index.

The exploratory moderator analysis found no baseline patient characteristic that robustly predicted a larger or smaller 120-day response. This null result should be interpreted cautiously rather than as evidence of true non-moderation: interaction effects generally require substantially larger samples than main effects to detect reliably, and with 50 participants this analysis was likely underpowered for anything but a large moderation effect. It was also limited to each outcome’s own baseline severity, since symptom chronicity and other clinical covariates were not available for this analysis; these remain plausible candidate moderators for future, adequately powered work.

From a clinical standpoint, these findings suggest that patients and clinicians should not expect the respiratory and chest-wall-mobility benefits of this technique to fade after treatment ends: on this evidence, the opposite pattern was observed. This has practical implications for how the technique is positioned within a physiotherapy plan of care — as an intervention whose value may become more, not less, apparent at longer-term review — while the corresponding absence of benefit for expiratory-phase mobility indicates that clinicians should not extrapolate this durability to every dimension of respiratory function.

This study has several limitations. First, the diaphragmatic technique was delivered on a fixed schedule (sessions 1, 3, 5, 6, 8 and 10) interspersed with conventional-physiotherapy-only sessions in the experimental group; the present analysis cannot isolate the contribution of any single session, and does not attempt to. Second, no home exercise programme or maintenance strategy was prescribed between the end of treatment and the 120-day assessment, and unmonitored self-directed activity during this period could contribute to the observed durability. Third, all participants were recruited from a single private clinic, which may limit the generalizability of these findings to other clinical settings or populations. Fourth, the moderator analysis, as noted above, was likely underpowered and restricted to baseline severity as the only available candidate moderator. Fifth, participants could not be blinded to group allocation given the hands-on nature of the intervention, and no sham diaphragmatic technique was used as a comparator; expectation effects cannot be ruled out, and confirmation in a sham-controlled design is warranted.

## 5. Conclusions

Adding manual diaphragmatic stretching to conventional physiotherapy for non-specific low back pain was associated with greater inspiratory thoracic expansion, and with higher FVC, FEV1 and peak expiratory flow measured with a portable spirometer, than conventional physiotherapy alone. These differences arose during the treatment period and were maintained 120 days after treatment ended. Expiratory-phase chest wall mobility and mid-to-late expiratory flows did not benefit. The size of the spirometric differences, the lack of participant blinding and of a sham comparator, mean that these findings need confirmation in sham-controlled trials using laboratory spirometry. No baseline characteristic moderated the long-term response, although this exploratory analysis was likely underpowered.

## Data Availability

The data underlying the results presented in this study cannot be shared publicly due to restrictions imposed by the informed consent signed by participants, which limits data use to the scope of the original study, and in accordance with the European General Data Protection Regulation (GDPR). These restrictions were reviewed and upheld by the Research Ethics Committee of the University of Seville, Spain (approval ref. 1032-N-23). Data access requests may be submitted to the Institutional Repository eIDUS of the University of Seville, which serves as the non-author institutional point of contact for data availability inquiries.

## Acknowledgements

The authors thank the participants for their involvement in the study.

## Suporting information

S1 CONSORT Checklist. CONSORT 2025 checklist.

S1 Protocol. Trial protocol approved by the Research Ethics Committee of the University of Seville (original and English translation).

S1 Appendix. Treatment manual and TIDieR checklist.

S1 Table. Mean (SD) of each outcome by group and assessment, with mean change from baseline to day 120.

S2 Table. Uncorrected and FDR-corrected p-values for all between-group comparisons.

S3 Table. Full results of the exploratory moderator analysis.

S4 Table. Sensitivity analyses of the 120-day estimates and individual-level FVC change.

## Notes

### Competing Interest Statement

The authors have declared no competing interest.

### Clinical Trial

ClinicalTrials.gov NCT06069388

### Author Declarations

The study was approved by the Research Ethics Committee of the University of Seville, Spain (ref. 1032-N-23). All participants provided written informed consent before enrolment. The study was conducted in accordance with the Declaration of Helsinki.

